# COVID-19 in Great Britain: epidemiological and clinical characteristics of the first few hundred (FF100) cases: a descriptive case series and case control analysis

**DOI:** 10.1101/2020.05.18.20086157

**Authors:** Nicola L Boddington, Andre Charlett, Suzanne Elgohari, Jemma L Walker, Helen I McDonald, Chloe Byers, Laura Coughlan, Tatiana Garcia Vilaplana, Rosie Whillock, Mary Sinnathamby, Nikolaos Panagiotopoulos, Louise Letley, Pauline MacDonald, Roberto Vivancos, Obaghe Edeghere, Joseph Shingleton, Emma Bennett, Daniel J Grint, Helen Strongman, Kathryn E Mansfield, Christopher Rentsch, Caroline Minassian, Ian J Douglas, Rohini Mathur, Maria Peppa, Simon Cottrell, Jim McMenamin, Maria Zambon, Mary Ramsay, Gavin Dabrera, Vanessa Saliba, Jamie Lopez Bernal

## Abstract

**Objectives:** Following detection of the first virologically-confirmed cases of COVID-19 in Great Britain, an enhanced surveillance study was initiated by Public Health England to describe the clinical presentation, course of disease and underlying health conditions associated with infection of the first few hundred cases.

**Methods:** Information was collected on the first COVID-19 cases according to the First Few X WHO protocol. Case-control analyses of the sensitivity, specificity and predictive value of symptoms and underlying health conditions associated with infection were conducted. Point prevalences of underlying health conditions among the UK general population were presented.

**Findings:** The majority of FF100 cases were imported (51.4%), of which the majority had recent travel to Italy (71.4%). 24.7% were secondary cases acquired mainly through household contact (40.4%). Children had lower odds of COVID-19 infection compared with the general population.

The clinical presentation of cases was dominated by cough, fever and fatigue. Non-linear relationships with age were observed for fever, and sensitivity and specificity of symptoms varied by age.

Conditions associated with higher odds of COVID-19 infection (after adjusting for age and sex) were chronic heart disease, immunosuppression and multimorbidity.

**Conclusion:** This study presents the first epidemiological and clinical summary of COVID-19 cases in Great Britain. The FFX study design enabled systematic data collection. The study characterized underlying health conditions associated with infection and set relative risks in context with population prevalence estimates. It also provides important evidence for generating case definitions to support public health risk assessment, clinical triage and diagnostic algorithms.

## Introduction

In December 2019, the World Health Organisation (WHO) was notified of a cluster of unexplained pneumonia cases detected in Wuhan city, China. This was subsequently identified and designated as COVID-19, caused by the novel severe acute respiratory syndrome coronavirus 2 (SARS-CoV-2), a strain previously unseen in humans. The UK was one of the first countries affected in Europe, with its first two confirmed cases of COVID-19 detected on 31 January 2020 (1).

The epidemiology and clinical features of early COVID-19 cases identified in China and elsewhere have been reported (2-7). In a pre-print article, Ma *et al*., present a pooled analysis of 1,155 cases from seven countries with estimates of key epidemiological parameters (8) and the first identified cases in the WHO European Region have been described (1). The most commonly reported symptoms have been fever, fatigue, dry cough, myalgia and dyspnoea (1-5, 8, 9). These studies did not report on the sensitivity, specificity or positive predictive values of symptoms.

Reported estimates of proportions of cases with underlying health conditions vary between 23% and 51% (2-5, 9). A range of underlying health conditions have been found to be associated with poor outcomes from COVID-19 (10-12). However, there are few controlled studies of underlying health conditions as risk factors for infection. In a pre-print from the United States, a test-negative design study of 3789 Veterans tested for COVID-19 found that the only comorbidity associated with testing positive after multivariable adjustment was chronic obstructive pulmonary disease, which was associated with decreased risk of testing positive for COVID-19(13). Studies in other settings and with general population controls (not selected by testing) are needed to understand the role of underlying health conditions in infection itself, rather than severity of illness.

The First Few X (FFX) is an established enhanced surveillance system designed to investigate the clinical and epidemiological characteristics of at least the first 100 confirmed cases of an emerging infectious disease and their close contacts (14). The design has previously been used in the 2009 influenza H1N1 pandemic (15) and the WHO has recommended that countries adopt this protocol for investigation of COVID-19 (14). Following the detection of the first GB cases at the end of January 2020, the FF100 surveillance system for COVID-19 was initiated by Public Health England (PHE). The primary objectives were to describe the clinical presentation and course of disease and underlying health conditions associated with for infection.

This paper describes the epidemiological and clinical characteristics of the first few hundred cases of COVID-19 identified in GB, including estimates of sensitivity and specificity of selected symptoms, associations of underlying health conditions with infection and estimates of the prevalence of these conditions in the UK population.

## Methods

After detection of COVID-19 in China, the FF100 protocol, data collection questionnaires and electronic data capture system were modified for the COVID-19 outbreak.

### ▪ Case ascertainment

Initially enhanced surveillance forms were collected for all virologically-confirmed cases of COVID-19 in GB. Due to the large predominance of imported cases during February 2020, this was later restricted to sporadic cases only. Case definitions for testing and the time periods that they applied are outlined in Table 1.

**Table 1:** Summary of possible case definitions for population testing

| Time period | Case definition |
| --- | --- |
| Before 7 February 2020 | **Epidemiological criteria**<br>In the 14 days before the onset of illness:<br>• travel to China<br>OR<br>• contact with confirmed cases of COVID-19 (previously referred to as 2019-nCoV infection)<br>**and**<br>**Clinical criteria**<br>• severe acute respiratory infection requiring admission to hospital with clinical or radiological evidence of pneumonia or acute respiratory distress syndrome<br>OR<br>• acute respiratory infection of any degree of severity, including at least one of shortness of breath (difficult breathing in children) or cough (with or without fever)<br>OR<br>• fever with no other symptoms |
| From 7 February 2020 | **Epidemiological criteria**<br>In the 14 days before the onset of illness:<br>• travel to affected countries [the list of affected countries expanded between 7 February and 13 March 2020]. This includes transit, for any length of time, in these countries<br>OR<br>• contact with confirmed cases of COVID-19 (previously referred to as 2019-nCoV infection)<br>**and**<br>**Clinical criteria**<br>• severe acute respiratory infection requiring admission to hospital with clinical or radiological evidence of pneumonia or acute respiratory distress syndrome<br>OR<br>• acute respiratory infection of any degree of severity, including at least one of shortness of breath (difficult breathing in children) or cough (with or without fever)<br>OR<br>• fever with no other symptoms |
| From 13 March 2020 | **Inpatient definition**<br>• requiring admission to hospital (a hospital practitioner has decided that admission to hospital is required with an expectation that the patient will need to stay at least one night)<br>**and**<br>• have either clinical or radiological evidence of pneumonia<br>**or**<br>• acute respiratory distress syndrome<br>**or**<br>• influenza like illness (fever $\geq 37.8^{\circ}\text{C}$ and at least one of the following respiratory symptoms, which must be of acute onset: persistent cough (with or without sputum), hoarseness, nasal discharge or congestion, shortness of breath, sore throat, wheezing, sneezing) |

Imported cases were defined as cases with travel to countries with known COVID-19 circulation at the time or with contact with a confirmed case whilst abroad within a maximum incubation period of their onset of symptoms. Secondary cases were defined as cases that had contact with a confirmed or probable/suspected case in the UK and did not fit the definition of an imported case. Sporadic cases were defined as cases with no travel history to countries with known COVID-19 circulation, and no known contact with a confirmed case.

### ▪ Data sources

#### ○ Case questionnaires

Data collection was coordinated by the PHE Surveillance Cell, National Infection Service.

On identification of a positive case, the local Health Protection Teams, Field Service Teams and the equivalents in the Devolved Administrations of Great Britain, were asked to complete an initial enhanced surveillance questionnaire (Form 1A) providing information on demographic details, medical history and travel history. This information was collected as soon as possible after a positive laboratory result was reported and was collected through interview with the cases and/or healthcare workers or family members. Cases were followed up after 14 days from initial report (Form 1B). Follow-up information on cases was collected to determine the occurrence of any medical complications and clinical outcome. To improve completeness of the initial questionnaires and to achieve a high rate of follow-up, a team of PHE health protection practitioners, nurses, doctors and field epidemiologists proactively followed up the FF100 cases in England using telephone interviews.

Data from completed forms were entered into a dedicated FF100 web-tool. Data were extracted, cleaned and quality checked.

#### ○ Control data

##### ▪ Possible cases who tested negative

During the early stages of the epidemic more limited data on patient demographics, presenting illness, clinical course/complications after onset and exposures in the 14 days before onset of first symptoms, were collected on all possible cases using a Minimum Data Set (MDS) form. Those suspected cases testing negative were used as a control group for the analyses of symptoms.

##### ▪ General population electronic health record data

General population data were obtained from an anonymised dataset of primary care electronic health records, Clinical Practice Research Datalink (CPRD) GOLD. In the UK, almost 99% of the population is registered at a general practice and healthcare is free at the point of delivery. CPRD is broadly representative of the population in terms of age and sex (16). Primary care prescriptions and laboratory test results are automatically recorded electronically. Diagnoses are recorded using Read codes, and generally have good positive predictive value (17). Approximately 75% of CPRD practices in England have consented to data linkage, and for patients at these practices anonymised secondary care data (Hospital Episode Statistics (HES)) is linked to primary care records.

### ▪ Data analysis

Descriptive analyses of the FF100 cases were undertaken in relation to patient characteristics, clinical symptoms and complications, healthcare interactions and outcomes. For the analysis of symptoms, missing data was assumed to indicate absence of that symptom. Ethnicity was assigned to cases by linking, using NHS number and Date of Birth, to the latest recording of ethnicity in the Outpatient HES database. If no record of ethnicity was found in the Outpatient record, then the HES Admitted Patient Care recording of ethnicity was used.

Underlying health conditions among the UK general population were ascertained from diagnoses, prescriptions and test results in CPRD primary care records, with hospital diagnoses supplementing ascertainment for a secondary analysis limited to England (due to availability of HES-linkage). Detailed definitions of risk groups were tailored to respiratory infection risk using established methods described more fully in Supplementary Table A (18-22). Multimorbidity was defined as ≥2 conditions in different domains, combining asthma with chronic respiratory disease and organ transplant with other immunosuppression.

Analysis of associations of underlying health conditions with infection used a case-control design, with controls from the UK general population in CPRD (details in Supplementary Table A). Cases with any data recorded for any underlying health condition (whether ‘Yes’ or ‘No’) or any medical history recorded in free text were included, with missing data assumed to indicate absence of that comorbidity. Cases for whom there was no evidence of any data collection for underlying health conditions were excluded (n=23). Cancer in controls was defined as any history of malignant cancer, as questionnaire free-text suggested this would be most comparable to cases.

Single variable and multivariable logistic regression were used to calculate odds ratios of each underlying health condition in COVID-19 cases compared to the UK general population, adjusted for sex and age (in categories <18, 18-35, 36-49, 50-69 and ≥70 years) as *a priori* confounders. Consideration of other potential confounders (such as comorbid health conditions) was limited by data sparsity. Age (over and under 70 years) and infection source (imported or sporadic/secondary) were considered as potential effect modifiers.

Point prevalence estimates for underlying health conditions were calculated per 100,000 for the UK general population on 5 March 2019. Prevalence of cancer was estimated for malignant cancer incident within the previous year rather than ever, for greater clinical relevance.

Estimates of the sensitivity and specificity of symptoms were obtained from the proportion of COVID-19 cases, and those testing negative respectively. Predictive values were estimated for the observed prevalence of positive patients. The functional relationships between the presence of a symptom and age were explored using locally weighted scatter plot smoothing. Fractional polynomial logistic regression models were used to obtain parametric functions of these relationships with age. Interaction terms between the fractional polynomial age terms and case type (imported, sporadic or secondary) were used to assess if there was evidence of different age relationships. Logistic regression analyses were performed to assess which symptoms were independently associated with COVID-19, accounting for sex and age. Multinomial regression models were used with case type as the outcome variable to assess whether the associations with symptoms differed for each case type.

Analyses were undertaken in Microsoft Excel 2010, R and Stata 16 MP.

### ▪ Ethical considerations

This was an observational surveillance system carried out under the permissions granted under Regulation 3 of The Health Service (Control of Patient Information) Regulations 2020, and without explicit patient permission under Section 251 of the NHS Act 2006.

Analysis involving CPRD data was approved by the Independent Scientific Advisory Group of the CPRD (ISAC, reference 20_062A) and the London School of Hygiene and Tropical Medicine Ethics Committee (reference 21815). The ISAC protocol was made available to reviewers.

## Results

381 confirmed cases of COVID-19 in GB were included in this analysis (up to 09 April 2020) (359 from England, 19 from Scotland and three from Wales). 16.8% of cases were resident in London. Figure 1 shows the distribution of cases included in the FF100 study by date of symptom onset and COVID-19 case types. Approximately half of the FF100 cases were imported (51.4%) with the remainder being secondary (24.7%) or sporadic (23.9%) cases. Of the FF100 cases, 38.3% reported travel to Italy in the 14 days prior to symptom onset (71.4% of imported cases) and hence Europe was the most commonly visited continent by the imported cases (Figure 2). Where occupation was recorded (n=357), 42 cases were healthcare workers (11.8%), although the majority of these were imported cases (n=26). Of the secondary cases, almost all reported close contact with a confirmed case (93/94, 98.9%), of these 39.8% had close contact within a household setting, 10.8% in a healthcare setting, 47.3% in other settings (for example work setting, social gatherings) and 3.2% unknown setting.

**Figure 1.**
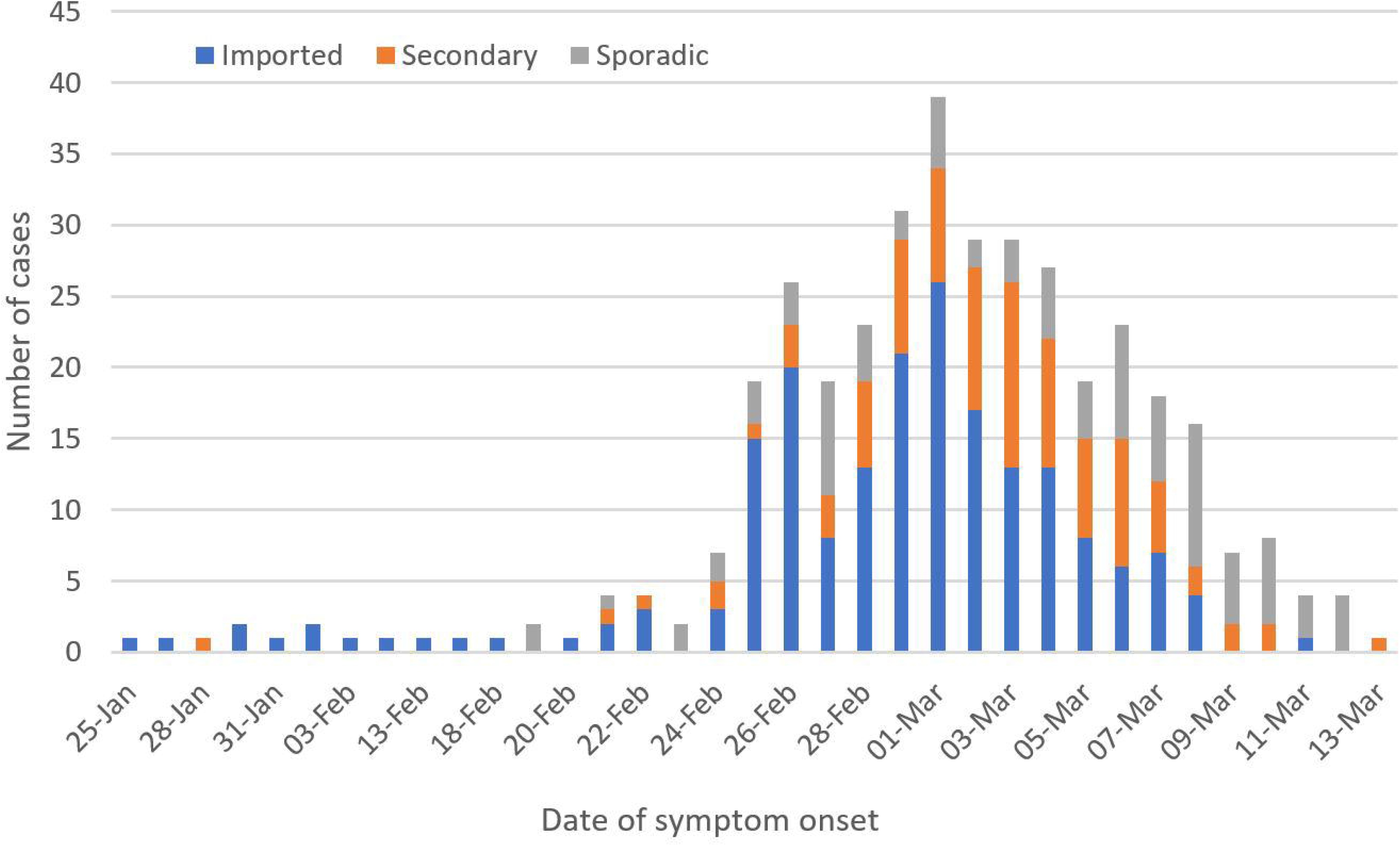

**Figure 2.**
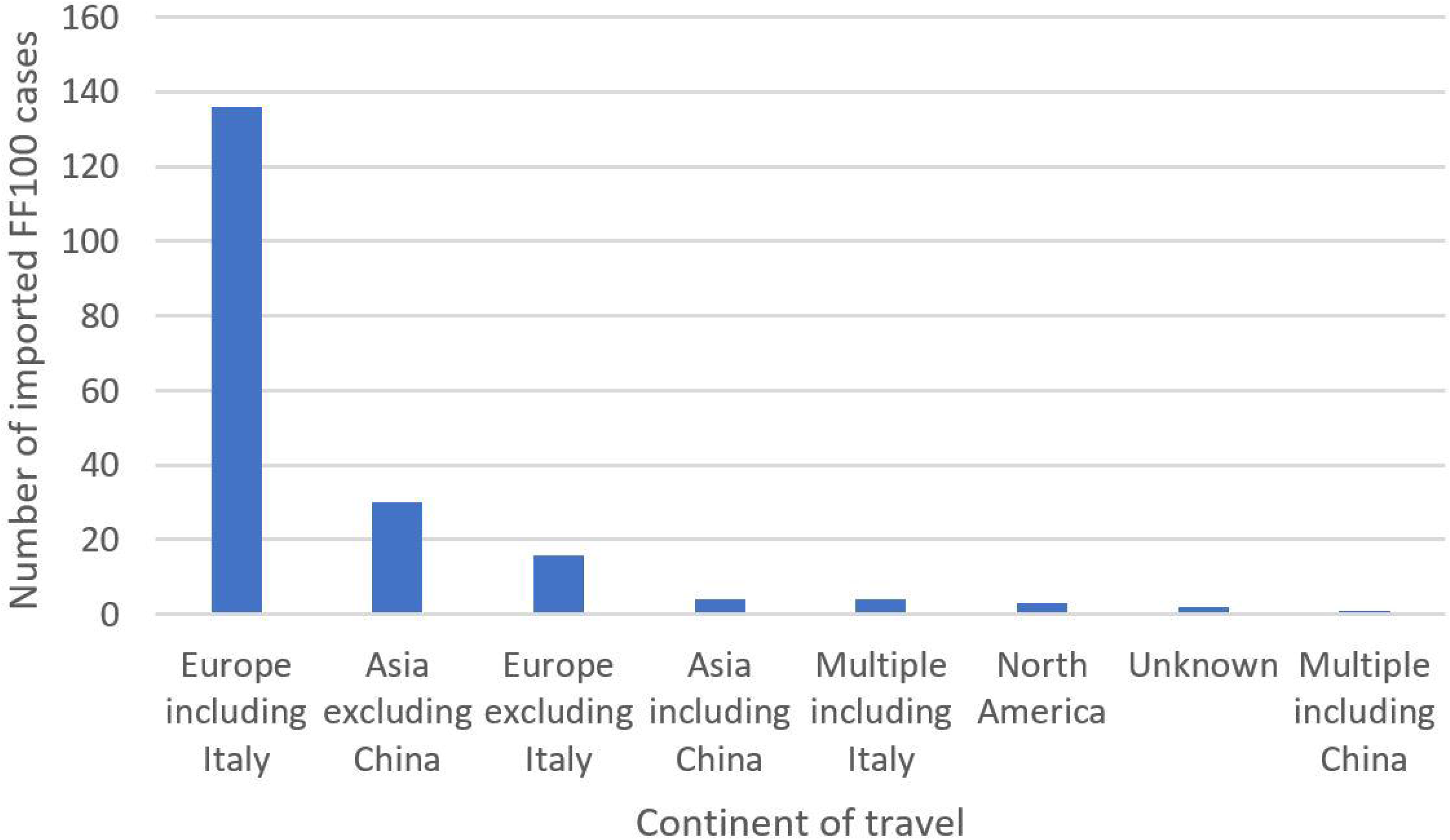

### ▪ Demographic characteristics of cases

There were slightly more male cases (56.7%) compared with female cases (43.3%). The FF100 cases ranged between 1 year and 94 years of age with a mean age of 47.7 years (standard deviation (SD) 17.4) (Figure 3). Country of birth was available on 68.2% of the cases, of which the majority were born in the UK (73.5%), followed by Italy (5.8%), Iran (1.9%) and the United States (1.9%). The ethnicity breakdown of the FF100 cases (available for 63.0% of cases) was comparable to the general population (England and Wales, ONS 2011 Census) (Table 2).

**Figure 3.**
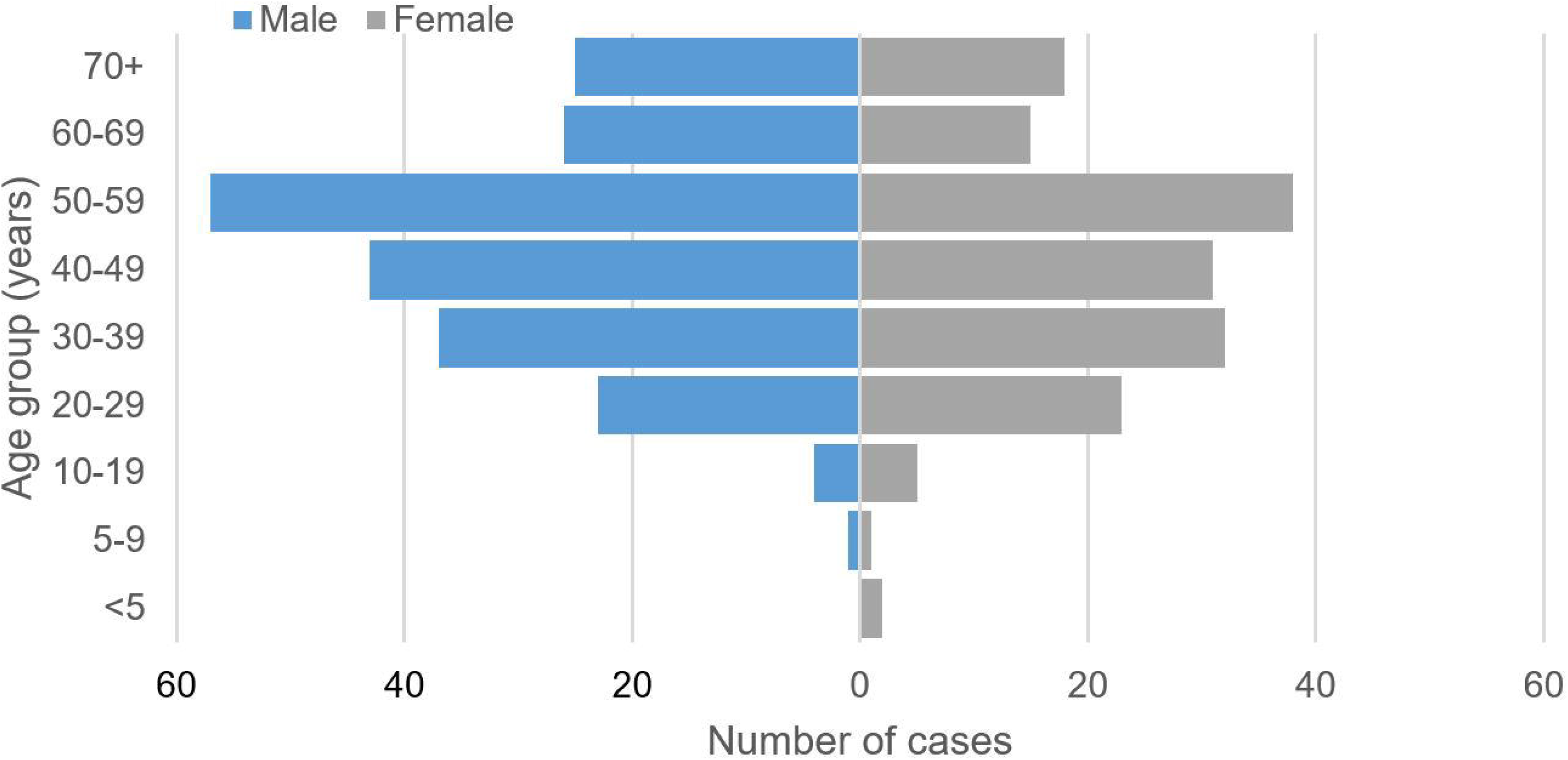

**Table 2:** Ethnicity of the FF100 cases compared to the general population (England and Wales)

| Ethnic group* | Number (%) | England & Wales (%)** |
| --- | --- | --- |
| White | 204 (85.0) | 86.0 |
| Asian / Asian British | 15 (6.3) | 7.5 |
| Black/African/ Caribbean/Black British | 9 (3.8) | 3.3 |
| Other ethnic group | 8 (3.3) | 1.0 |
| Mixed | 4 (1.7) | 2.2 |

\*Office for National Statistics (ONS) ethnic categories, \*\*From the 2011 ONS Census (https://www.ethnicity-facts-figures.service.gov.uk/uk-population-by-ethnicity/national-and-regional-populations/population-of-england-and-wales/latest)

Logistic regression analysis of associations of age and sex with COVID-19 included 358 cases with data on underlying health conditions (to allow adjustment for immunosuppression), and 2,705,963 UK general population controls.

When stratified by infection source and sex, women had lower odds of imported COVID-19 than men (AOR 0.61, 95% CI 0.46-0.83), but there was no evidence of an association of sex with secondary/sporadic cases (Table 3).

When stratified by infection source and age, children aged under 18 years had considerably lower odds of imported and secondary/sporadic COVID-19 than the general population (Table 2). Adults aged 50-69 years had higher odds of imported COVID-19 than adults aged 18-35 years, but otherwise there was no evidence of any association between infection and age for adults aged under 70 years. Adults aged 70 years or over had lower odds of imported COVID-19 than younger adults, but this association was not observed among secondary/sporadic cases (Table 3).

#### ○ Underlying health conditions

Analysis of associations of underlying health conditions with COVID-19 infection included 358 cases with comorbidity data, and 2,705,963 UK general population controls (Table 4). Approximately one third of the cases had an underlying health condition (115/358, 32.1%), and 11.7% of cases had multimorbidity. The most frequent conditions were asthma, diabetes, and chronic heart disease.

After adjustment for age and sex, cases had higher odds of chronic heart disease (adjusted odds ratio (AOR) 1. 95, 95% CI 1.34–2.84), immunosuppression (AOR 4.46, 2.65–7.50), and multimorbidity (AOR 1.68, 1.17–2.40), and lower odds of chronic kidney disease (AOR 0.39, 0.21–0.73) compared to the general population. There was weak evidence suggesting that cases had higher odds of chronic liver disease (AOR 2.66, 0.99–7.14) and lower odds of asthma (AOR 0.73, 0.53–1.02) than the general population.

When stratified by infection source, associations between underlying health conditions and COVID-19 infection appeared generally smaller for imported cases than for sporadic or secondary cases, but confidence intervals overlapped for all conditions other than multimorbidity.

There was no evidence of any association of COVID-19 infection with diabetes, chronic respiratory disease (excluding asthma), chronic neurological disease, organ transplant or a history of malignant cancer, although confidence intervals were particularly wide for organ transplant.

In England, age-and sex-adjusted odds of liver disease (AOR 3.79, 95% CI 1.41–10.18) and organ transplant (2.64, 9.37–18.80) were higher among cases than the general population, but no evidence of these associations remained in analysis using linked hospital data among patients eligible for data linkage (Supplementary Table B). When secondary care data were used to enhance ascertainment of underlying health conditions in England, the increased ascertainment of underlying health conditions among controls resulted in a general decrease in odds ratios for associations of those conditions with COVID-19 infection – this was most pronounced for chronic liver disease, immunosuppression and chronic heart disease. Nevertheless, odds of chronic heart disease and immunosuppression remained consistently higher among cases than the general population in all analyses.

**Table 3:**
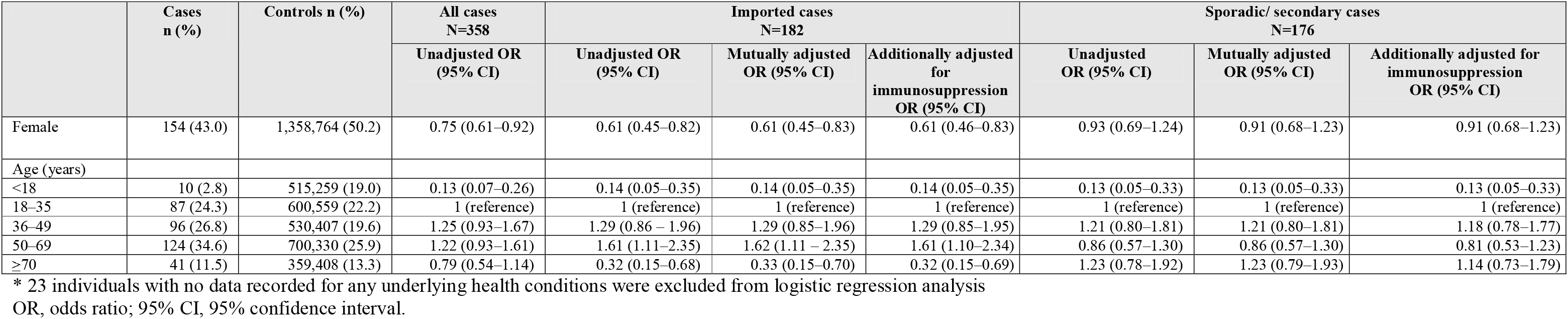
Associations between age, sex and COVID-19 infection, for the first few hundred GB cases (N=358)* compared to the UK general population (N=2,705,963)

|  | Cases<br>n (%) | Controls n (%) | All cases<br>N=358<br>Unadjusted OR<br>(95% CI) | Imported cases<br>N=182 |  |  | Sporadic/ secondary cases<br>N=176 |  |  |
| --- | --- | --- | --- | --- | --- | --- | --- | --- | --- |
|  |  |  |  | Unadjusted OR<br>(95% CI) | Mutually adjusted<br>OR (95% CI) | Additionally adjusted<br>for<br>immunosuppression<br>OR (95% CI) | Unadjusted<br>OR (95% CI) | Mutually adjusted<br>OR (95% CI) | Additionally adjusted for<br>immunosuppression<br>OR (95% CI) |
| Female | 154 (43.0) | 1,358,764 (50.2) | 0.75 (0.61–0.92) | 0.61 (0.45–0.82) | 0.61 (0.45–0.83) | 0.61 (0.46–0.83) | 0.93 (0.69–1.24) | 0.91 (0.68–1.23) | 0.91 (0.68–1.23) |
| Age (years) |  |  |  |  |  |  |  |  |  |
| <18 | 10 (2.8) | 515,259 (19.0) | 0.13 (0.07–0.26) | 0.14 (0.05–0.35) | 0.14 (0.05–0.35) | 0.14 (0.05–0.35) | 0.13 (0.05–0.33) | 0.13 (0.05–0.33) | 0.13 (0.05–0.33) |
| 18–35 | 87 (24.3) | 600,559 (22.2) | 1 (reference) | 1 (reference) | 1 (reference) | 1 (reference) | 1 (reference) | 1 (reference) | 1 (reference) |
| 36–49 | 96 (26.8) | 530,407 (19.6) | 1.25 (0.93–1.67) | 1.29 (0.86 – 1.96) | 1.29 (0.85–1.96) | 1.29 (0.85–1.95) | 1.21 (0.80–1.81) | 1.21 (0.80–1.81) | 1.18 (0.78–1.77) |
| 50–69 | 124 (34.6) | 700,330 (25.9) | 1.22 (0.93–1.61) | 1.61 (1.11–2.35) | 1.62 (1.11 – 2.35) | 1.61 (1.10–2.34) | 0.86 (0.57–1.30) | 0.86 (0.57–1.30) | 0.81 (0.53–1.23) |
| ≥70 | 41 (11.5) | 359,408 (13.3) | 0.79 (0.54–1.14) | 0.32 (0.15–0.68) | 0.33 (0.15–0.70) | 0.32 (0.15–0.69) | 1.23 (0.78–1.92) | 1.23 (0.79–1.93) | 1.14 (0.73–1.79) |

\* 23 individuals with no data recorded for any underlying health conditions were excluded from logistic regression analysis
OR, odds ratio; 95% CI, 95% confidence interval.

**Table 4:** Associations between underlying health conditions and COVID-19 infection, for the first few hundred GB cases (N=358)* compared to the UK general population (N=2,705,963)o Prevalence of underlying health conditions in the general population

|  | Cases<br>n (%) | Controls<br>n (%) | Crude odds ratio<br>OR (95% CI) | Adjusted for age<br>and sex<br>AOR (95% CI) | P value | Imported cases<br>N=182<br>AOR (95% CI) | Sporadic/ secondary<br>cases<br>N=176<br>AOR (95% CI) | Age <70 years<br>N=317 cases,<br>2,346,555 controls<br>AOR (95% CI) | Age ≥70 years<br>N=41 cases, 359,408<br>controls<br>AOR (95% CI) |
| --- | --- | --- | --- | --- | --- | --- | --- | --- | --- |
| Chronic liver disease | 4 (1.1) | 9,481 (0.4) | 3.21 (1.20–8.61) | 2.66 (0.99–7.14) | 0.05 | 1.26 (0.18–8.99) | 4.22 (1.34–13.25) | 3.25 (1.21–8.75) | NA |
| Chronic heart disease | 30 (8.4) | 121,041 (4.5) | 1.95 (1.34–2.84) | 1.95 (1.30–2.93) | 0.001 | 1.27 (0.63–2.56) | 2.62 (1.57–4.39) | 1.58 (0.89–2.79) | 2.54 (1.36–4.76) |
| Chronic respiratory<br>disease | 17(4.7) | 87,387 (3.2) | 1.49 (0.92–2.43) | 1.39 (0.84–2.30) | 0.20 | 0.52 (0.16–1.64) | 2.23 (1.26–3.97) | 1.21 (0.62–2.36) | 1.70 (0.78–3.68) |
| History of asthma (ever) | 40 (11.2) | 377,443 (13.9) | 0.78 (0.56–1.08) | 0.73 (0.53–1.02) | 0.07 | 0.77 (0.49–1.22) | 0.70 (0.43–1.12) | 0.69 (0.48–0.99) | 1.15 (0.48–2.73) |
| Chronic neurological<br>disease | 15 (4.2) | 87,698 (3.2) | 1.31 (0.78–2.19) | 1.24 (0.73–2.11) | 0.43 | 0.76 (0.28–2.06) | 1.64 (0.87–3.11) | 1.33 (0.68–2.60) | 1.09 (0.46–2.60) |
| Diabetes mellitus | 35 (9.8) | 192,779 (7.1) | 1.41 (1.00–2.00) | 1.23 (0.85–1.77) | 0.26 | 0.53 (0.26–1.09) | 2.06 (1.33–3.19) | 0.88 (0.54–1.42) | 2.47 (1.32–4.61) |
| Organ transplant<br>recipient | 1 (0.3) | 2,477 (0.1) | 3.06 (0.43–21.8) | 2.47 (0.35–17.6) | 0.37 | NA | 5.45 (0.76–39.00) | 2.79 (0.39–19.93) | NA |
| Immunosuppression | 15 (4.2) | 22,836 (0.8) | 5.14 (3.06–8.62) | 4.46 (2.65–7.50) | <0.001 | 1.72 (0.55–5.38) | 7.47 (4.13–13.48) | 4.42 (2.48–7.89) | 4.58 (1.41–14.85) |
| Chronic kidney disease | 12 (3.4) | 195,315 (7.2) | 0.45 (0.25–0.79) | 0.39 (0.21–0.73) | 0.002 | 0.09 (0.01–0.68) | 0.57 (0.29–1.12) | 0.42 (0.16–1.14) | 0.39 (0.18–0.84) |
| History of cancer (ever) | 17 (1.1) | 128,183 (4.7) | 1.00 (0.62–1.63) | 0.93 (0.56–1.54) | 0.77 | 0.49 (0.18–1.33) | 1.32 (0.73–2.39) | 1.03 (0.56–1.89) | 0.76 (0.32–1.81) |
| Multimorbidity | 42 (11.7) | 204,925 (7.6) | 1.62 (1.18–2.24) | 1.68 (1.17–2.40) | 0.004 | 0.75 (0.38–1.51) | 2.76 (1.76–4.32) | 1.38 (0.96–2.22) | 2.37 (1.28–4.39) |
| Multimorbidity (without<br>asthma)* | 36 (10.1) | 168,118 (6.2) | 1.69 (1.20–2.38) | 1.83 (1.24–2.71) | 0.002 | 0.65 (0.28–1.52) | 3.20 (1.97–5.19) | 1.36 (0.77–2.41)) | 2.69 (1.45–4.98) |

OR, odds ratio; AOR, odds ratio adjusted for age and sex; 95% CI, 95% confidence interval.
\* 23 individuals with no data recorded for any underlying health conditions were excluded from logistic regression analysis
Adjusted odds ratios were adjusted for age (under 18 years, 18–35 years, 36–49 years, 50–69 years, and ≥70 years) and sex.
Multimorbidity was defined as two or more of the following: chronic liver disease; chronic heart disease; chronic respiratory disease (including asthma); chronic neurological disease; immunosuppression (including organ transplant recipient), or chronic kidney disease. A previous history of cancer was not included in estimates of multimorbidity in case-control analysis. As a *post hoc* analysis considering the possibility of over ascertainment of multimorbidity among controls by including all individuals with a history of asthma, the multimorbidity analysis was repeated additionally excluding asthma.

#### ○ Prevalence of underlying health conditions in the general population

The prevalence of any underlying health condition was 26.7% in England, 28.8% in Scotland, 30.4% in Northern Ireland, and 30.8% in Wales (Supplementary Table C). Among those conditions associated with COVID-19, point prevalence of chronic heart disease ranged from 3.7% (England) to 5.0% (Scotland), immunosuppression from 0.7% (Wales) to 1.0% (Scotland), chronic liver disease from 0.3% (England) to 0.5% (Scotland) and multimorbidity from 6.6% (England) to 8.4% (Wales and Northern Ireland). Prevalence of any underlying health condition increased to 30.0% in England when ascertainment was supplemented with secondary care data; the greatest increases in prevalence estimates were for chronic liver disease and chronic heart disease.

### ▪ Clinical features

#### ○ Symptoms over the course of illness

The symptoms during illness reported by all FF100 cases are summarised in Figure 4. The most frequent symptoms were cough (77.7%), fatigue (70.9%), fever (60.1%), headache (56.7%), muscle ache (50.9%), appetite loss (44.1%). Just over three quarters of the cases experiencing a cough reported whether they experienced a dry or productive cough: the majority of whom reported a dry cough (78.1%). Anosmia was added to the follow-up questionnaire part way through the FF100 study. 111 of the 229 cases who were asked this question reported anosmia during their illness (48.5%). One case reported anosmia as their only symptom.

Cough was the most common presenting symptom for all age groups. A lower proportion of cases in the 70+ year old age group reported headache, sore throat, runny nose and sneezing compared with other age groups (Supplementary Table D).

**Figure 4.**
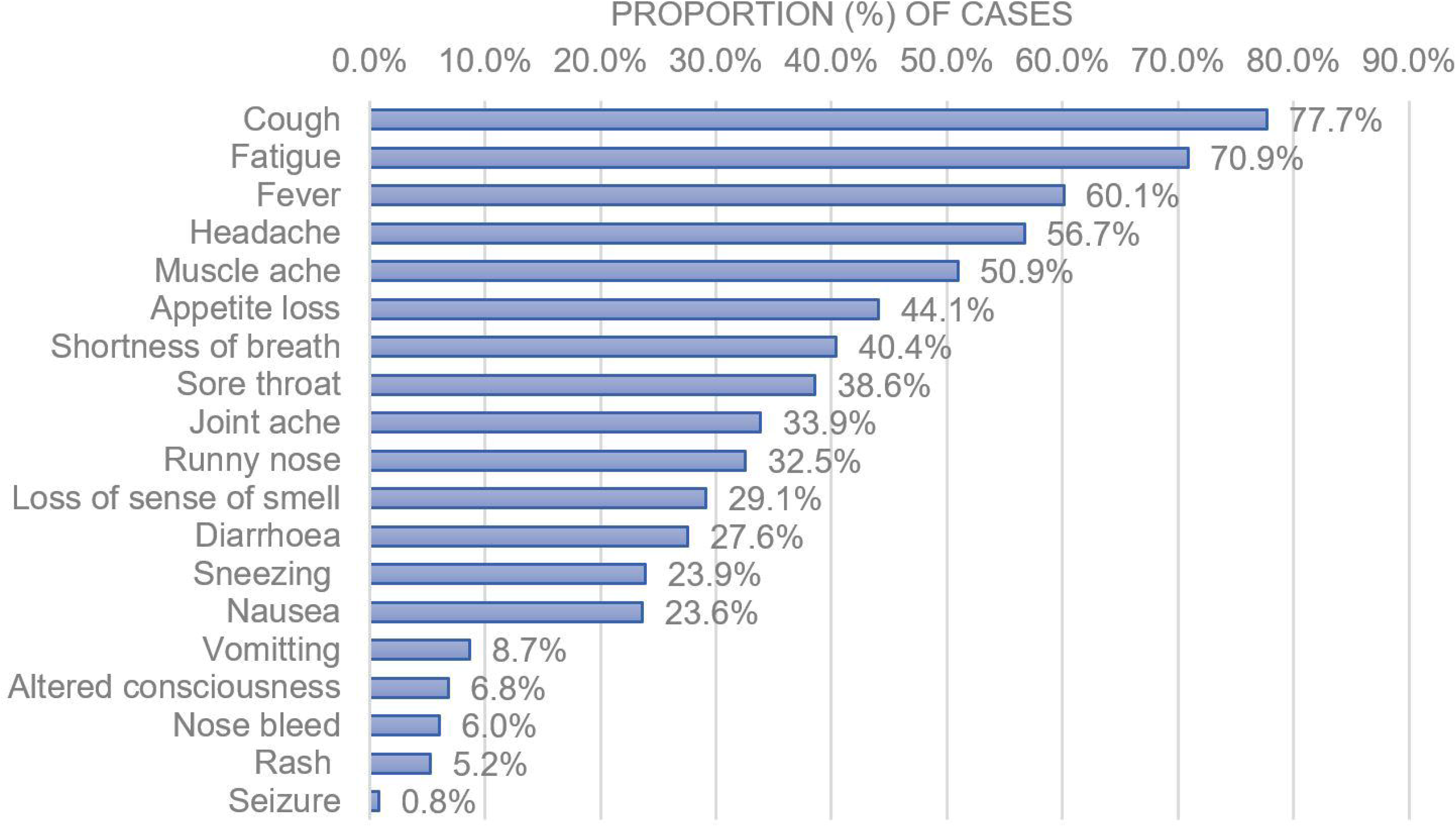

Symptoms by sex were relatively consistent, although a higher proportion of females reported headache (61.6% compared to 51.6% of males), sore throat (43.9% compared to 33.2% of males), joint ache (43.9% compared to 33.2% of males), diarrhoea (32.3% compared to 22.1% of males) and nausea (30.5% compared to 16.6% in males) (Supplementary Table D). Table 5 shows common groups (pairs/trios) of presenting symptoms.

**Table 5:** Common groups of symptoms amongst FF100 cases

|  | **Number of cases (%)** | **Imported (%)** | **Sporadic (%)** | **Secondary (%)\*** |
| --- | --- | --- | --- | --- |
| **Pairs of symptoms** |  |  |  |  |
| Cough, fatigue | 216 (56.7) | 106 (54.1) | 60 (65.9) | 50 (53.2) |
| Cough, fever | 185 (48.6) | 83 (42.3) | 64 (70.3) | 38 (40.4) |
| Fatigue, headache | 177 (46.5) | 101 (51.5) | 31 (34.1) | 45 (47.9) |
| Fever, fatigue | 174 (45.7) | 80 (40.8) | 55 (60.4) | 39 (41.5) |
| Fatigue, muscle ache | 172 (45.1) | 91 (46.4) | 43 (47.3) | 38 (40.4) |
| Cough, headache | 168 (44.1) | 93 (47.4) | 32 (35.2) | 43 (45.7) |
| Cough, muscle ache | 156 (40.9) | 77 (39.3) | 41 (45.1) | 38 (40.4) |
| Fatigue, appetite loss | 149 (39.1) | 65 (33.2) | 50 (54.9) | 34 (36.2) |
| **Trios of symptoms** |  |  |  |  |
| Fever, cough, fatigue | 142 (37.3) | 60 (30.6) | 51 (56.0) | 31 (33.0) |
| Cough, fatigue, headache | 141 (37.0) | 76 (38.8) | 28 (30.8) | 37 (39.4) |
| Cough, fatigue, muscle ache | 139 (36.5) | 69 (35.2) | 38 (41.8) | 32 (34.0) |
| Cough, fatigue, appetite loss | 124 (32.5) | 49 (25.0) | 45 (49.5) | 30 (31.9) |
| Fatigue, muscle ache, headache | 122 (32.0) | 68 (34.7) | 25 (27.5) | 29 (30.9) |
| Fever, fatigue, muscle ache | 115 (30.2) | 57 (29.1) | 34 (37.4) | 24 (25.5) |
| Fever, fatigue, appetite loss | 113 (29.7) | 46 (23.5) | 43 (47.3) | 24 (25.5) |
| Fatigue, muscle ache, joint ache | 110 (28.9) | 56 (28.6) | 31 (34.1) | 23 (24.5) |
| Cough, shortness of breath, fatigue | 107 (28.1) | 40 (20.4) | 39 (42.9) | 28 (29.8) |
| Fever, cough, appetite loss | 103 (27.0) | 39 (19.9) | 42 (46.2) | 22 (23.4) |

\*1 secondary case was asymptomatic so was excluded

### ▪ Association of symptoms with COVID-19

There were four symptoms; fever, cough, shortness of breath and sore throat, that were collected on 380 symptomatic COVID-19 cases in the FF100, and 752 contemporaneous confirmed non-COVID-19 respiratory infections.

The relationship between the presence of a symptom and age for those with COVID-19 and those with non-COVID-19 respiratory illness is shown in Figures 5 and 6. These show non-linear relationships with age. For fever, there is an increase in occurrence with increasing age for COVID-19, while for those with other respiratory infections the occurrence decreases with increasing age.

**Figure 5.**
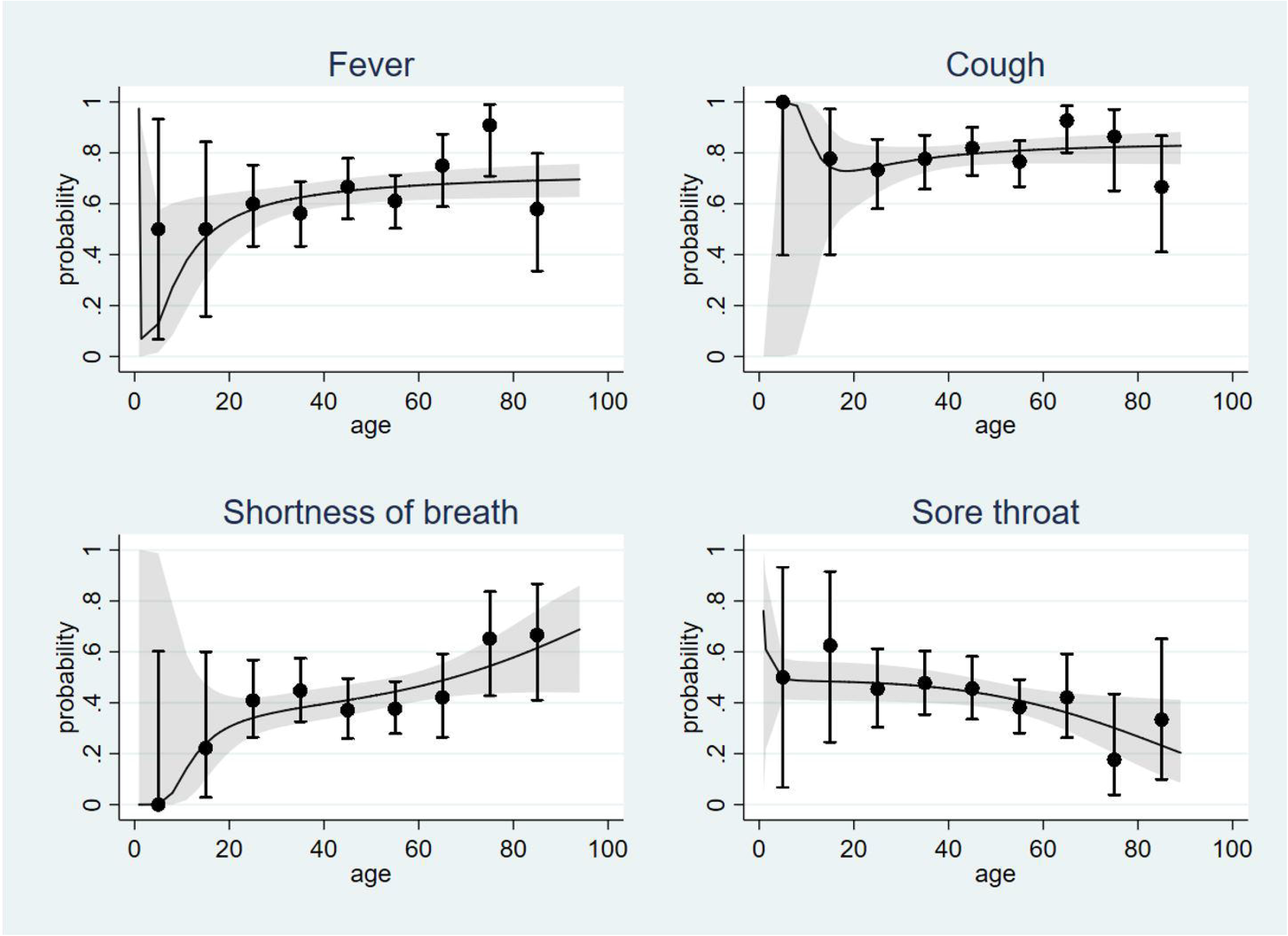

**Figure 6.**
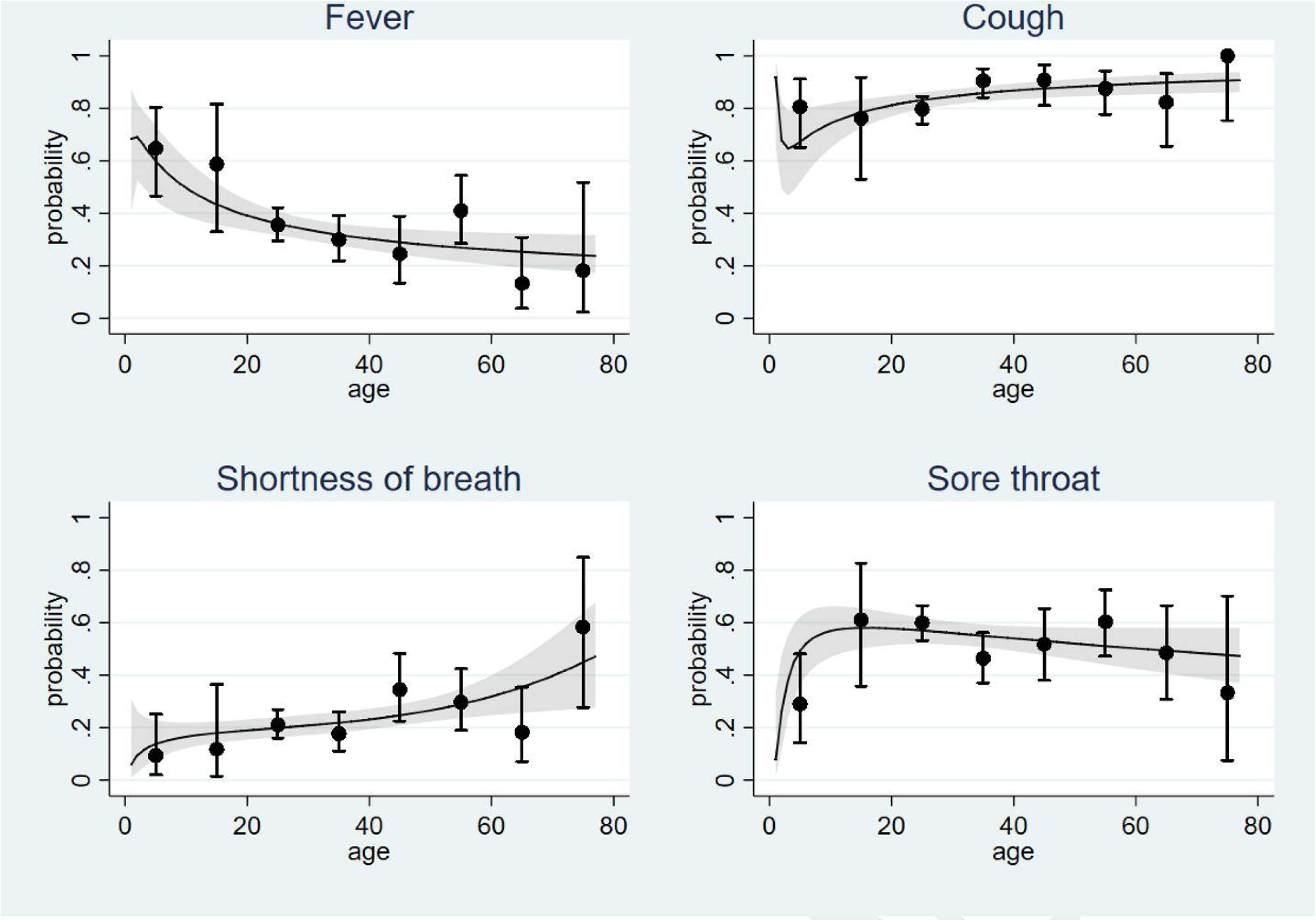

For cough, a similar relationship is observed with COVID-19 and other respiratory infections. Shortness of breath exhibits an increasing occurrence with increasing age for both, although in those with COVID-19 there is a higher proportion of elderly cases with this symptom compared to those with other respiratory infections. The age relationship for sore throat was similar for both COVID-19 and other respiratory infections.

The shape of the age relationship with each symptom were similar in each of the different types of COVID-19 cases (imported, sporadic and secondary).

#### ○ Sensitivity and specificity

Estimates of sensitivity and specificity for symptoms are presented in Table 6. Broad age categories; <30, 30-59, 60+, have been used to provide estimates within these age strata. Fever had both good sensitivity and specificity (64.0% and 63.9%), cough had high sensitivity but poor specificity (79.6% and 15.5%), shortness of breath was the most specific symptom but had low sensitivity, while sore throat had relatively low sensitivity and specificity.

**Table 6:** Sensitivity and specificity of symptoms (95% CI)

| Estimate | Fever | Cough | Shortness of breath | Sore throat | Fever and/or cough | Fever and cough |
| --- | --- | --- | --- | --- | --- | --- |
| **Sensitivity** |  |  |  |  |  |  |
| All subjects | 64.0%<br>(58.8%-69.0%) | 79.6%<br>(75.1%-83.6%) | 42.1%<br>(37.0%-47.3%) | 42.4%<br>(37.1%-47.8%) | 90.9%<br>(87.5%-93.6%) | 48.2% (43.0%-53.3%) |
| Imported cases | 61.0%<br>(53.4%-68.2%) | 77.2%<br>(70.6%-82.9%) | 28.3%<br>(22.0%-35.2%) | 41.9%<br>(34.8%-49.4%) | 90.7%<br>(85.7%-94.4%) | 42.1% (35.0%-49.3%) |
| Sporadic cases | 83.7%<br>(74.2%-90.8%) | 87.2%<br>(78.3%-93.4%) | 67.8%<br>(56.9%-77.4%) | 37.8%<br>(26.8%-49.9%) | 93.2%<br>(85.7%-97.5%) | 71.4% (61.0%-80.4%) |
| Secondary cases | 77.4%<br>(67.6%-85.4%) | 46.6%<br>(35.9%-57.5%) | 47.1%<br>(36.3%-58.1%) | 89.1%<br>(80.9%-94.7%) | 77.4%<br>(67.6%-85.4%) | 38.3% (28.5%-48.9%) |
| Below 30 years of age | 57.7%<br>(43.2%-71.3%) | 75.9%<br>(62.8%-86.1%) | 35.1%<br>(22.9%-48.9%) | 48.2%<br>(34.7%-62.0%) | 94.7%<br>(85.4%-98.9%) | 33.9% (22.1%-47.4%) |
| 30 to 59 years of age | 61.4%<br>(54.6%-67.8%) | 78.5%<br>(72.7%-83.6%) | 39.6%<br>(33.2%-46.2%) | 43.3%<br>(36.7%-50.1%) | 90.2%<br>(85.6%-93.7%) | 45.1% (38.7%-51.7%) |
| Aged 60 or older | 75.3%<br>(64.5%-84.2%) | 85.2%<br>(75.6%-92.1%) | 54.4%<br>(42.8%-65.7%) | 34.3%<br>(23.2%-46.9%) | 90.2%<br>(81.7%-95.7%) | 66.7% (55.5%-76.6%) |
| **Specificity** |  |  |  |  |  |  |
| All subjects | 63.9%<br>(60.1%-67.6%) | 15.5%<br>(13.0%-18.3%) | 75.5%<br>(72.1%-78.8%) | 46.2%<br>(42.3%-50.1%) | 8.3%<br>(6.4%-10.5%) | 76.2% (73.0%-79.2%) |
| Below 30 years of age | 59.5%<br>(53.5%-65.3%) | 20.5%<br>(16.1%-25.4%) | 80.9%<br>(75.7%-85.3%) | 43.5%<br>(37.5%-49.6%) | 10.1%<br>(7.0%-14.1%) | 74.5% (69.3%-79.3%) |
| 30 to 59 years of age | 68.3%<br>(61.8%-74.3%) | 10.2%<br>(6.9%-14.5%) | 74.9%<br>(68.8%-80.3%) | 48.5%<br>(41.9%-55.1%) | 5.6%<br>(3.2%-9.1%) | 78.4% (72.9%-83.1%) |
| Aged 60 or older | 85.4%<br>(70.8%-94.4%) | 12.8%<br>(4.8%-25.7%) | 71.1%<br>(55.7%-83.6%) | 54.8%<br>(38.7%-70.2%) | 12.8%<br>(4.8%-25.7%) | 87.2% (74.3%-95.2%) |

#### ○ Predictive values

Estimates of the positive predictive value (PPV) and negative predictive value (NPV) for the observed proportions of cases are presented in Table 7. The PPV and NPV were similar for fever and shortness of breath. These were 48.9% and 76.7%, respectively, for fever and 48.7% and 70.2% for shortness of breath, respectively. The estimated PPV increased with increasing age groups, with the NPV decreasing.

**Table 7:** Predictive values of presenting symptoms

| Estimate | Fever | Cough | Shortness of breath | Sore throat | Fever and/or cough | Fever and cough |
| --- | --- | --- | --- | --- | --- | --- |
| **All subjects** |  |  |  |  |  |  |
| PPV | 48.9%<br>(44.3%-53.6%) | 32.3%<br>(29.3%-35.4%) | 48.7%<br>(43.1%-54.4%) | 29.8%<br>(25.8%-34.1%) | 33.3%<br>(30.4%-36.6%) | 50.6%<br>(45.3%-55.8%) |
| NPV | 76.7%<br>(72.9%-80.2%) | 60.0%<br>(52.7%-67.0%) | 70.2%<br>(66.7%-73.6%) | 59.8%<br>(55.3%-64.1%) | 64.2%<br>(53.7%-73.8%) | 74.4%<br>(71.2%-77.5%) |
| **Imported COVID-19 cases** |  |  |  |  |  |  |
| PPV | 31.4%<br>(26.5%-36.6%) | 19.4%<br>(16.6%-22.3%) | 25.0%<br>(19.4%-31.3%) | 18.4%<br>(14.8%-22.4%) | 20.5%<br>(17.9%-23.4%) | 31.4%<br>(25.8%-37.4%) |
| NPV | 85.8%<br>(82.4%-88.8%) | 72.2%<br>(64.5%-79.0%) | 78.5%<br>(75.1%-81.6%) | 73.3%<br>(68.7%-77.6%) | 77.2%<br>(66.4%-85.9%) | 83.5%<br>(80.5%-86.2%) |
| **Sporadic COVID-19 cases** |  |  |  |  |  |  |
| PPV | 23.4%<br>(18.8%-28.5%) | 10.8%<br>(8.6%-13.3%) | 26.7%<br>(21.0%- 33%) | 7.49%<br>(5.0%-10.6%) | 10.8%<br>(8.7%-13.2%) | 26.6%<br>(21.2%-32.7%) |
| NPV | 96.8%<br>(94.6%-98.2%) | 91.2%<br>(84.8%-95.5%) | 94.7%<br>(92.4%-96.4%) | 86.6%<br>(82.5%- 90.0%) | 91.0%<br>(81.5%-96.6%) | 95.7%<br>(93.7%-97.1%) |
| **Secondary COVID-19 cases** |  |  |  |  |  |  |
| PPV | 16.3%<br>(12.2%-21.2%) | 10.4%<br>(8.2%-12.9%) | 20.2%<br>(14.9%-26.4%) | 10.6%<br>(7.7%-14.1%) | 10.8%<br>(8.7%-13.2%) | 16.7%<br>(12.0%-22.4%) |
| NPV | 90.5%<br>(87.4%-93.0%) | 84.4%<br>(77.2%-90.1%) | 91.4%<br>(88.7%-93.6%) | 86.6%<br>(82.5%-90.0%) | 85.9%<br>(75.6%-93.0%) | 90.8%<br>(88.3%-92.9%) |
| **Below 30 years of age** |  |  |  |  |  |  |
| PPV | 21.0%<br>(14.6%-28.6%) | 15.4%<br>(11.4%-20.2%) | 27.4%<br>(17.6%-39.1%) | 15.1%<br>(10.2%-21.2%) | 16.4%<br>(12.6%-20.9%) | 20.2%<br>(12.8%-29.5%) |
| NPV | 88.3%<br>(82.8%-92.5%) | 81.6%<br>(71.0%-89.5%) | 85.8%<br>*81.0%-89.8% | 80.1%<br>(72.2%-86.3%) | 91.2%<br>(76.3%-98.1%) | 85.6%<br>(80.0%-89.5%) |
| **30 to 59 years of age** |  |  |  |  |  |  |
| PPV | 65.2%<br>(58.3%-71.7%) | 43.6%<br>(38.8%-48.5%) | 60.7%<br>(52.4%-68.5%) | 44.9%<br>(38.2%-51.8%) | 45.7%<br>(41.1%-50.3%) | 64.8%<br>(57.0%-72.1%) |
| NPV | 64.6%<br>(58.2%-70.6%) | 35.1%<br>(24.5%-46.8%) | 55.9%<br>(50.2%-61.4%) | 46.9%<br>(40.4%-53.4%) | 39.5%<br>(24.0%-56.6%) | 61.8%<br>(56.4%-67.0%) |
| **Aged 60 or older** |  |  |  |  |  |  |
| PPV | 91.0%<br>(81.5%-96.6%) | 62.7%<br>(53.0%-71.8%) | 76.8%<br>(63.6%-87.0%) | 54.8%<br>(38.7%-70.2%) | 64.3%<br>(54.9%-73.1%) | 90.3%<br>(80.1%-96.4%) |
| NPV | 63.6%<br>(49.6%-76.2%) | 33.3%<br>(13.3%-59.0%) | 47.1%<br>(34.8%-59.6%) | 34.3%<br>(23.2%-46.9%) | 42.9%<br>(17.7%-71.1%) | 59.4%<br>(46.9%-71.1%) |

#### ○ Multivariable analysis of presenting symptoms and diagnosis of COVID-19

The association between COVID-19 and symptoms has been assessed using logistic regression modelling. The estimated odds ratios (OR) and adjusted odds ratios (AOR) results are presented in Table 8.

**Table 8:** Association between COVID-19 and symptoms, age and sex

| Symptom | Single variable analysis |  | Multivariable analysis* |  |
| --- | --- | --- | --- | --- |
|  | OR | 95% CI (p value) | AOR | 95% CI (p value) |
| Fever | 3.15 | 2.41-4.13 (<0.001) | 4.15 | 2.95-5.82 (<0.001) |
| Cough | 0.71 | 0.52-0.99 (0.04) | 0.73 | 0.48-1.09 (0.13) |
| Shortness of breath | 2.24 | 1.71-2.95 (<0.001) | 2.27 | 1.56-3.29 (<0.001) |
| Sore throat | 0.63 | 0.48-0.82 (0.001) | 0.78 | 0.56-1.10 (0.16) |
| Age (10 years) | 1.65 | 1.52- 1.79 (<0.001) | 1.63 | 1.47-1.81 (<0.001) |
| Male | 1.45 | 1.13-1.85 (0.004) | 1.26 | 0.90-1.76 (0.19) |

\*adjusted for other symptoms, age and sex

After adjusting for the other symptoms, age and sex; fever and shortness of breath were significantly associated with a diagnosis of COVID-19 (AOR 4.15 (95% CI 2.95-5.82) and (AOR 2.27 (95% CI 1.56-3.29), respectively. Cough and sore throat exhibited no association with COVID-19. Age was positively associated which may reflect how the early cases were identified rather than what may have be occurring in the general population.

As the occurrence of symptoms change with age, interaction terms between a symptom and the broad age groups used previously have been explored. This provided strong evidence that the association with fever differed in these age groups (interaction term p value 0.0009). In the under 30-year olds the AOR was 2.67 (95% CI 1.41-5.07), in the 30 to 59-year old the AOR was 4.08 (95% CI 2.60-6.41), while in those 60 years of age or older the AOR was 17.15 (95% CI 5.60-52.55). No other symptom exhibited any strong evidence of an interaction with age.

Finally, a multinomial regression model was used to assess associations with the different COVID-19 case types. As with the logistic regression models, there was strong evidence of an interaction with age group and fever (Table 9). Sporadic cases exhibit a stronger association with both fever and shortness of breath, and have a higher proportion aged 60 or older. For all case types the association with fever is strongest in those aged 60 or older. Cough and sore throat are not strongly associated with any case type.

**Table 9:** Estimated ARRR (95% CI) of COVID-19 for different symptoms and demographic characteristics from a multinomial logistic regression model

| Predictor | Imported | Sporadic | Secondary |
| --- | --- | --- | --- |
| **Symptoms** |  |  |  |
| Fever (in <30 year olds) | 3.35 (1.48-7.58) | 5.86 (0.59-58.03) | 1.04 (0.36-3.01) |
| Fever (30-59 year olds) | 3.38 (2.02-5.65) | 15.62 (6.08-40.16) | 3.01 (1.59-5.71) |
| Fever (60+ years old) | 19.61 (5.57-69.08) | 17.54 (4.42-69.55) | 10.30 (2.08-51.05) |
| Cough | 0.53 (0.33-0.86) | 0.88 (0.39-1.97) | 0.61 (0.33-1.12) |
| Shortness of breath | 1.64 (1.03-2.58) | 5.36 (2.92-9.82) | 2.78 (1.64-4.27) |
| Sore throat | 0.76 (0.51-1.14) | 0.60 (0.33-1.10) | 0.97 (0.58-1.60) |
| **Age group** |  |  |  |
| Aged less than 30 | Reference | Reference | reference |
| 30 to 59 year olds | 5.06 (2.45-10.47) | 6.45 (0.76-54.87) | 2.90 (1.33-6.32) |
| 60+ year olds | 3.29 (1.19-9.07) | 23.78 (2.71-208.8) | 1.74 (0.50-5.98) |
| **Sex** |  |  |  |
| Male | 1.50 (1.00-2.23) | 1.17 (0.64-2.12) | 0.78 (0.47-1.30) |

### ▪ Healthcare interactions, clinical course and complications

Overall follow-up information was obtained on 88.7% of FF100 cases (338/381) (94.2% of the FF100 cases in England (338/359)).

Duration of illness, in those cases which had sufficient information recorded (n=154), ranged from 2 to 36 days (median 11 days, IQR 7-15 days, mean 12.1 days).

The use of NHS 111, a telephone and online service was the most common healthcare interaction amongst the FF100 cases in England with just over three quarters of cases accessing NHS 111 at least once (Table 9). Smaller proportions of cases visited their GP (13.1%) or Accident and Emergency (A&E) (28.7%). 42.9% of cases were hospitalised of which 35 were admitted to ICU (21.5% of those hospitalised) and 25 required mechanical ventilation (15.3% of those hospitalised) (Table 10). Clinical outcomes were ascertained for 302 of these cases. At the time of follow-up 72.8% of cases had recovered from their COVID-19 illness (220/302), 18.9% were still ill (self-reported illness) (57/302) and 8.3% had died (25/302).

**Table 10:** Healthcare interactions and clinical course of the FF100 cases in England

|  | NHS111 (%) | GP visit (%) | A&E visit (%) | Hospitalised (%) | Admitted to ICU (%) | Mechanical ventilation (%) | ARDS* (%) | Chest X-ray evidence of pneumonia | ECMO** |
| --- | --- | --- | --- | --- | --- | --- | --- | --- | --- |
| **FF100 cases in England (n=359)** | 277 (77.2) | 47 (13.1) | 103 (28.7) | 154 (42.9) | 35 (9.7) | 25 (7.0) | 12 (3.3) | 57 (15.9) | 1 (0.3) |

## Discussion

This study presents an early assessment of the epidemiological and clinical characteristics of COVID-19 patients in GB. The COVID-19 FF100 system was rapidly established with data collected and analysed from the first virologically-confirmed cases. In this early phase, patients were only eligible for testing if they fitted the case definition at the time, which was based on a combination of clinical symptoms, travel history or contact with a confirmed case and later admission to hospital. This is the first epidemiological and clinical summary of severe and non-severe confirmed COVID-19 cases in GB and is also the first controlled analysis that we are aware of outside the United States that investigates associations of underlying health conditions with COVID-19 infection, rather than severe outcomes. The study is also the first to estimate sensitivity and specificity of different symptoms for COVID-19 in the GB population.

Just over half of the cases included in our study were imported with the majority imported from Italy, highlighting the importance of the Italian outbreak in facilitating spread to other European countries (23). Sporadic cases were detected within a month of the first confirmed case in GB.

Where known exposure to a confirmed case was documented, the most common setting for exposure was the household, confirming the household as a high risk setting for transmission. Only a small proportion of the cases were healthcare workers and most of these acquired their infection abroad suggesting that, in the containment phase of the epidemic, healthcare acquired infection did not play a significant role.

There were slightly more males amongst the FF100 cases than females; a gender distribution that has been described elsewhere (2-5). However, there was no evidence of an association of sex with secondary/sporadic cases. The median age was 47.7 years, again similar to other case series (2-5, 8, 9). Children had lower odds of infection compared to the general population for all case types.

The proportion of cases with any underlying health condition was 32.1%, slightly lower than the 37.6% of 7,162 cases reported to CDC in the US (24). Chronic heart disease, immunosuppression and multimorbidity were associated with higher odds of COVID-19 infection, for sporadic and imported cases. There was also weak evidence suggesting that chronic liver disease might be associated with higher odds of COVID-19 infection. These conditions may be risk factors for infection, although the associations could alternatively be explained by a greater likelihood of symptomatic or severe presentations in these patients which in turn increased the probability of testing for COVID-19.

The prevalence of any specified underlying health condition among the general population on 5 March 2019 ranged from 27.7% in England to 32.1% in Wales, similar to the proportion seen in the FF100 cases (32.1%). Chronic kidney disease and a history of asthma were associated with lower odds of COVID-19 infection for imported and sporadic cases, suggesting this is not explained by reduced likelihood of travel. This could be explained if ascertainment of these conditions is higher using electronic primary care records for general population controls than case questionnaires. For asthma this is plausible since the case questionnaires only asked about asthma requiring medication – although free text responses include mild or childhood asthma. It is also possible that asthma is not a risk factor for COVID-19 infection: asthma has been under-represented in case series of hospitalised patients, and there are mixed findings on the association of asthma with severe outcomes of COVID-19 (5, 11-13, 24-27).

The clinical presentation in FF100 cases was dominated by cough, fatigue and fever, consistent with other studies (2-5, 7-9). The proportion of FF100 cases reporting fever was slightly lower than case series descriptions from China (2-5), although similar to the first cases of COVID-19 described in Europe (1). This may be due to the inclusion of non-hospitalised cases in this study as well as the European study. Almost half of cases reported anosmia during the course of their illness, an unexpected symptom that has been observed by others, with some suggestion that it might precede the onset of more common symptoms or be present in those with mild or asymptomatic infection (28-31). Further work is required to understand anosmia as a distinctive clinical feature of COVID-19 (30, 31).

Symptoms were relatively consistent by sex. We noted differences in symptom presentation by age, with a lower proportion of cases in the youngest (<20 years) and oldest age groups (70+ years) reporting symptoms when compared to the other age groups. In particular, a non-linear relationship with age was observed for fever which increased with age for the COVID-19 cases. Although this study included only a small number of children, our findings are congruent with other studies suggesting that children may experience milder illness with different symptoms (32-34). The different sensitivity and specificity of symptoms by age highlights the need to consider this when setting up case definitions to support public health risk assessment, clinical triage and diagnostic algorithms.

Fever is clearly an important symptom, exhibiting good sensitivity and specificity. It was also significantly associated with a diagnosis of COVID-19, as was shortness of breath, and there was evidence of an interaction between fever and age, including for the different case types. In particular, sporadic cases exhibited a stronger association with fever and shortness of breath. The model used broad age groups, so differences due to age may have been concealed by this.

As expected, the non-urgent medical telephone and online service, NHS 111, was the most common healthcare interaction of the FF100 cases, in keeping with key government messaging during the study period which emphasised that those experiencing COVID-19 symptoms should stay home and contact NHS 111. This highlights the importance of public messaging about using online and telephone services to avoid propagating transmission of COVID-19 in healthcare settings. Forty-three percent of cases included in our study were hospitalised however this is certainly an overestimate of the case hospitalisation rate since at the beginning of the UK COVID-19 incident response all confirmed cases were hospitalised for isolation rather than clinical management purposes, and from 13 March 2020, testing was restricted to hospitalised cases only. The fact that only a small proportion of those hospitalised required mechanical ventilation in comparison with other studies supports this (2, 4).

One of the key strengths of this study is the detailed epidemiological and clinical data it was able to obtain. Use of the FFX study design allowed for systematic data collection on cases using ready to use infrastructure. The study achieved high rates of follow up on the FF100 cases and also benefits from having followed the majority of cases up past the intended 14 days. This has enabled capture of totality of symptoms over the course of illness which in many cases has been longer than 14 days.

This study included detailed ascertainment of underlying health conditions in controls from the general population to describe age and sex adjusted associations of health conditions with infection.

There were a number of limitations to this study. Firstly, the FF100 cases are a small subset of cases that were identified at the beginning of the COVID-19 epidemic in the Great Britain. They are unlikely to be representative of all GB cases of COVID-19 or of the general population, with different exposure and risk profiles. As such, the case-control study is vulnerable to selection bias. The FF100 cases are likely to be biased towards more severe cases who presented to healthcare, therefore they will underrepresent those with mild illness in the population, which may overestimate the associations between underlying health conditions and infection. The clinical presentation of FF100 cases may also differ to that of all GB cases since the imported cases (accounting for more than half of all cases) are more likely to have been of working age and may have been ‘healthier’ than the general population. This could conversely bias our estimates of associations between health conditions and infection downwards. The severity estimates are likely to be overestimates due to policy changes over the course of the study period, with hospitalisation of cases for isolation purposes initially and latterly restricting testing to hospitalised cases only.

Despite having a high follow-up rate, clinical outcome was not available on some cases due to sensitivities and difficulties around ascertaining the required information, particularly for the most severe cases. Also, for some cases the timing of follow-up, during which the initial questionnaire was revisited, may have been some time after onset of illness leading to possible recall bias.

The power and precision of estimates of associations between underlying health conditions and COVID-19 is limited by the number of cases in the FF100. This also limits the scope to adjust for potential confounders other than age and sex in the study. It is important to note that the predictive values do not necessarily reflect the prevalence of COVID-19 in the population at any particular time.

Future pandemic planning, should note the importance of maintaining FFX studies into the community transmission phase, as we have shown differences between imported and GB-acquired cases. Furthermore, achieving high quality and complete data capture and follow-up of cases, depends on the ability to rapidly mobilise a cadre of trained public health professionals. This can pose a challenge when capacity is already overwhelmed by the incident response. Consideration should also be given to case ascertainment through FFX investigations and how this may differ as a pandemic progresses due to changing contact tracing and testing policies over time.

In summary, this paper presents the first epidemiological and clinical summary of COVID-19 cases in Great Britain. This study has informed modelling of likely disease burden and healthcare demand, as well as priority groups for preventive measures such as physical distancing. This study also provides important evidence for generating case definitions to support public health risk assessment, clinical triage and diagnostic algorithms.

## Data Availability

Applications for relevant anonymised data should be submitted to the Public Health England Office for Data Release: https://www.gov.uk/government/publications/accessing-public-health-england-data/about-the-phe-odr-and-accessing-data

## Acknowledgements

We also acknowledge the contributions of Richard Pebody, Katie Owens, Hongxin Zhao and Chinelo Obi from the PHE Immunisation Department for their work on developing the protocols, study design, data collection and data entry, and the PHE Software Development Unit for their work on developing the FF100 web-platform.

We would like to acknowledge the contribution of a number of teams to this study including the PHE Health Protection Teams, PHE Field Services, PHE Centres, the PHE Centres and Regions Operational Cell, and the team of PHE health protection and immunisation specialist nurses and public health registrars who assisted with the case follow ups. Also the contribution of the teams in Health Protection Scotland, and Malorie Perry, Clare Sawyer and the staff of Public Health Communicable Disease Surveillance Centre and COVID-19 response cells in Public Health Wales.

We would also like to acknowledge the contributions of Nick Andrews, Anthony Scott, Julia Stowe, Liam Smeeth from the NIHR Health Protection Research Unit in Immunisation at PHE and LSHTM, and Angel Wong from LSHTM, towards study design, data extraction and cleaning, and interpretation of results for the analysis of underlying health conditions in cases and CPRD controls.

## Contributor information

JLB is the corresponding author and guarantor. NLB, AC, HIMcD, VS and JLB drafted the manuscript.

For the FF100 case description and analysis of symptoms: CB, LC, TV, RW, MS, and NP were involved in database management, data validation and supervision of data collection process; NLB, AC, JS and EB were involved in data analysis; LL, PMacD, RV, OE, SC, JMcM, VS, GD, JLB were involved in supervision of the investigation/data collection process.

For the analysis of underlying health conditions: HIMcD, JLW, HS, and ID designed the study, JLW, DJG, HS, KEM, CTR, CM, RM and MP conducted data analysis, HIMcD, JLW, DJG, HS, KEM, CTR, CM, ID, RM and MP interpreted the results, and HIMcD is the guarantor.

MR, MZ, JLB, SE, NLB and AC were involved in the conceptualization of the overall study. All authors reviewed and revised the manuscript and approved the final version.

## Data sharing

Applications for relevant anonymised data should be submitted to the Public Health England Office for Data Release: https://www.gov.uk/government/publications/accessing-public-health-england-data/about-the-phe-odr-and-accessing-data. The data used for this study were obtained from the Clinical Practice Research Datalink (CPRD). All CPRD data are available via an application to the Independent Scientific Advisory Committee (see https://www.cprd.com/Data-access). Data acquisition is associated with a fee.

